# Addressing Barriers to Interprofessional Working with Homecare Workers in Community Palliative Care: Insights from a Multi-site Qualitative Case Study in England

**DOI:** 10.1101/2025.08.14.25333668

**Authors:** Z Bayley, C Forward, C White, HL Elliott-Button, J Krygier, L Walker, M Pearson, J Hussein, PM Taylor, J Wray, A Roberts, H Bravington, MJ Johnson

## Abstract

**Background:** Social homecare workers are crucial in the provision of end-of-life care but are not part of the healthcare multidisciplinary team. Little is known about why they are excluded within interprofessional working practices.

**Aim:** To explore experiences of delivering and receiving end-of-life homecare, from multiple perspectives including HCWs and managers, people receiving care, carers, and social and healthcare practitioners.

**Design:** A qualitative multiple case study adopting a unique approach across three diverse sites using semi-structured interviews, and the option to create a Pictor chart – a visual diagram of relationships between those involved in care provision. Data were analysed using a reflexive thematic analysis. An adaptation of Bronfenbrenner’s ecological theory was used to inform the analysis.

**Setting/Participants:** 133 participants, were recruited from three economically and culturally different geographic areas within England.

**Results:** Although examples of good practice were seen, common barriers to collaboration between other practitioners and homecare workers were also identified. These included: lack of healthcare practitioner training on homecare workers’ role and its value/importance, lack of direct communication systems, gatekeeping of communication by managers, asynchronous working practices, one-sided communication, and restricted access to respective documentation and systems.

**Conclusion:** The homecare worker role was often poorly understood, undervalued, and with inadequate communication and interaction between practitioners, potentially impacting on quality of care. Collaborative practice is necessary for continuity of provision of high-quality care, but our findings indicate this was often absent due to knowledge, professional, and organisational barriers. Further research should explore suggested strategies to address the barriers identified.

**What is already known about this topic?:**

- Homecare workers provide a crucial service to people receiving care at end-of-life and wishing to remain in their own homes
- Collaborative working between social and healthcare services improves care provision.

**What this paper adds:**

- Barriers to communication led to ineffective collaborative working, including lack of direct communication systems with gatekeeping by managers, asynchronous working practices, misperception of reactive one-sided communication, and restricted access to respective documentation.
- Healthcare practitioners often had poor knowledge of and undervalued the homecare worker role, exacerbating poor communication and collaboration.
- Examples of good practice and potential solutions to barriers to interprofessional working

**Implications for practice, theory or policy:**

- Healthcare practitioners require training to better understand, recognise and appreciate the role of homecare workers
- Communication barriers should be addressed to enable the homecare worker to contribute effectively to the multi-disciplinary team.
- The homecare workforce’s role in providing high quality end-of-life care will remain constrained without wider professional and societal acknowledgement of its value.

## Background

Multi-disciplinary teams are foundational for the effective delivery of holistic end-of-life care^1,2^. Interprofessional working is needed with different professions working together with open communication, respect, and joint decision-making^3^. Despite global differences in homecare service delivery models homecare workers will benefit from effective interprofessional working as part of any multidisciplinary team delivering end-of life care. However, integrated care across and within care settings is challenging and varies across regions and settings. This can result in service provision which may not address the needs of people receiving care and cause marginalisation of some care workers^4,5^. This divide becomes problematic across health and social care in systems such as in the UK due to working within separate funding structures and organisational systems.

Within the UK, integration of services is a defining characteristic of the government’s long-term health plan^6^. The homecare worker role has not always been included in the rhetoric surrounding the integration of social and healthcare, for example in multidisciplinary meetings^7-9^. In practice, homecare workers usually operate in isolation from the rest of the community-based healthcare team and are often perceived as low-paid and low-skilled workers^10^.

## Methods

We conducted a wider study exploring the experiences, training and support needs of homecare workers providing care for people approaching end-of-life. We conducted a multiple case qualitative study across three regions in England and interviewed 133 stakeholders including people receiving homecare and their families, homecare workers, homecare managers, social care professionals and health practitioners involved in the delivery of community-based care at end-of-life. The protocol^11^ and other study findings are reported elsewhere (manuscripts in preparation).

In this paper we present findings in relation to interprofessional working from the themes generated from the full dataset.

### Theoretical framework and study design

We adopted the Bronfenbrenner’s human ecology theory wherein the homecare worker was seen as the primary person within an ecological environment^13^. We used an adapted version^14^ with levels in the environmental system differentiated based on their immediacy to the homecare worker, including time (chronosystem), those directly involved with care (microsystem), the interactions between those caregivers (mesosystem), the organisations relating to those caregivers (macrosystem), and the wider services involved such as government, the health service, and the third sector (exosystem). Social constructionism provided the theoretical underpinning evidenced in the co-creation of knowledge and meaning from participants.

Our design used the 3-stage Design of Case Study Research in Health Care (DESCARTE) case study model^14^ across three geographic sites in England. This enabled regional differences to be accounted for, supporting robust transferability^15^.

### Recruitment, sampling strategy and justification

Eligible participants were consenting adults (over 18 years), able to communicate in English or through an interpreter, with experience of home-based end-of-life care (defined as care within last 6 months of expected life) within the last 12 months. All participants were provided with information sheets and the opportunity to ask questions prior to giving either written or verbal informed consent.

Participants were recruited using purposive sampling as a strategy to maximise representation of different experiences of managing, delivering, and receiving care. The sampling framework was based on participant category (homecare worker, homecare worker manager, social or healthcare practitioner, service commissioner, person receiving care, informal carer) and region. Recruitment was carried out through homecare agencies, hospices, local NHS networks, health trusts, National Institute for Health and Care Research Regional Research Delivery Networks in each area, social media, snowballing, and the research team’s personal networks.

Our total planned sample size was 150, with the greatest proportion being homecare workers and homecare worker managers as the study’s primary focus. The sample size was guided by the concept of information power, and was informed by the study aim of recruiting sufficient numbers to allow sample diversity and quality^16^.

### Data generation

Interviews were conducted between May 2023 and May 2024 by experienced qualitative researchers, digitally recorded, transcribed verbatim, checked for accuracy by the researchers responsible for the interviews, and anonymised. A semi-structured topic guide (see Supplement 1), developed from the literature, research team, and service user and homecare worker advisory groups guided the interviews. The guide changed iteratively allowing unforeseen topics to be raised by participants and subsequently included. Participants were asked to describe their experiences of home-based care at end-of-life including a focus on training and support needs for homecare workers.

Participants could opt to create a Pictor diagram during the interview process. Pictor is a graphic elicitation technique using arrows to represent professional and social networks when working on a task^17^.

### Ethics

This study was registered on the Research Registry (No.8613) and was approved by the Health Research Authority (HRA) West Midlands Research Ethics Committee on 31^st^ March 2023 ref 23/WM/0030.

### Data analysis

Analysis was conducted using a reflexive thematic approach, adopting the six phases outlined by Braun and Clarke^18^. This enabled a deeper engagement with the data alongside an appreciation of the different perceptions and understandings of the participants and the research team. Data were organised using nVivo14 software in preparation for analysis. The research team worked collectively using reflexive practice including note-taking and open discussions to help reduce bias. Researchers familiarised themselves with the data by re-reading transcripts and where required, listening to audio recordings. The team compared and collected codes inductively from the data which were further developed and examined to form themes. Sense checking was carried out with the wider research team including our service user, family carer, and HCW advisors to ensure quality and validity of the analysis^19^. Pictor diagrams were analysed separately to the interviews and findings then synthesised narratively.

## Results

We interviewed 133 participants online or face-to-face between May 2023 – May 2024, at times and locations convenient to participants. Interviews varied in length, with two outliers of 7 minutes and 100 minutes. Forty-one Pictor diagrams were also created.

Most interviews were individual (n = 126), but five people receiving care expressed a preference for a family carer or homecare worker to join in a dyadic interview. Table 2 summarises participant characteristics. Identifier codes for illustrative quotations in are: site number (1, 2 or 3), type of participant (e.g. HOMECARE WORKER), and number of interview, for example 1-HOMECARE WORKER-4.

**Table 1:** Summary of participant characteristics.

| **Characteristics**<br>* x3 participants chose not to disclose | **Category** | **Total** |
| --- | --- | --- |
| **Age** | 18-20 | 1 |
|  | 21-30 | 12 |
|  | 31-40 | 21 |
|  | 41-50 | 27 |
|  | 51-60 | 31 |
|  | 61-70 | 19 |
|  | 71-80 | 8 |
|  | 81-90 | 7 |
|  | 91+ | 2 |
| **Self-reported gender** | Male | 27 |
|  | Female | 101 |
| **Self-reported ethnicity** | White British | 101 |
|  | Black British | 5 |
|  | Asian/ Asian British | 6 |
|  | Other White | 3 |
|  | Mixed | 4 |
|  | Other | 9 |
| **Participant Category** | **Identifier** |  |
| Homecare Worker | HOMECARE WORKER | 41 |
| Homecare Manager | MANAGER | 22 |
| Person receiving homecare | CLIENT | 18 |
| Family Carer | CARER | 20 |
| Practitioners including Service commissioners, nurses, General Practitioners*, social workers, and other healthcare practitioners | PRACTITIONER | 32 |

\*Family Doctor

The study generated 5 rich themes which can be found in Supplement B. This paper focuses on the identified theme of professional collaboration.

### Interprofessional collaboration throughout the eco-system

The theme of barriers to interprofessional collaboration was strongly generated from across the dataset. We found all the environmental systems had an impact on this issue (Figure 1). This included; the value and worth of homecare workers (microsystem), the direct interaction and communication between practitioners and homecare workers (mesosystem), the impact of structures and systems within the NHS, local authority, and charitable/private sector (exosystem), the wider societal impact of the lack of understanding and valuing of the homecare worker role (macrosystem), and the changing needs and influences of time on homecare workers, people receiving care, carers, and practitioners, such as asynchronous visits (chronosystem). Findings were consistent across all three sites.

**Figure 1:**
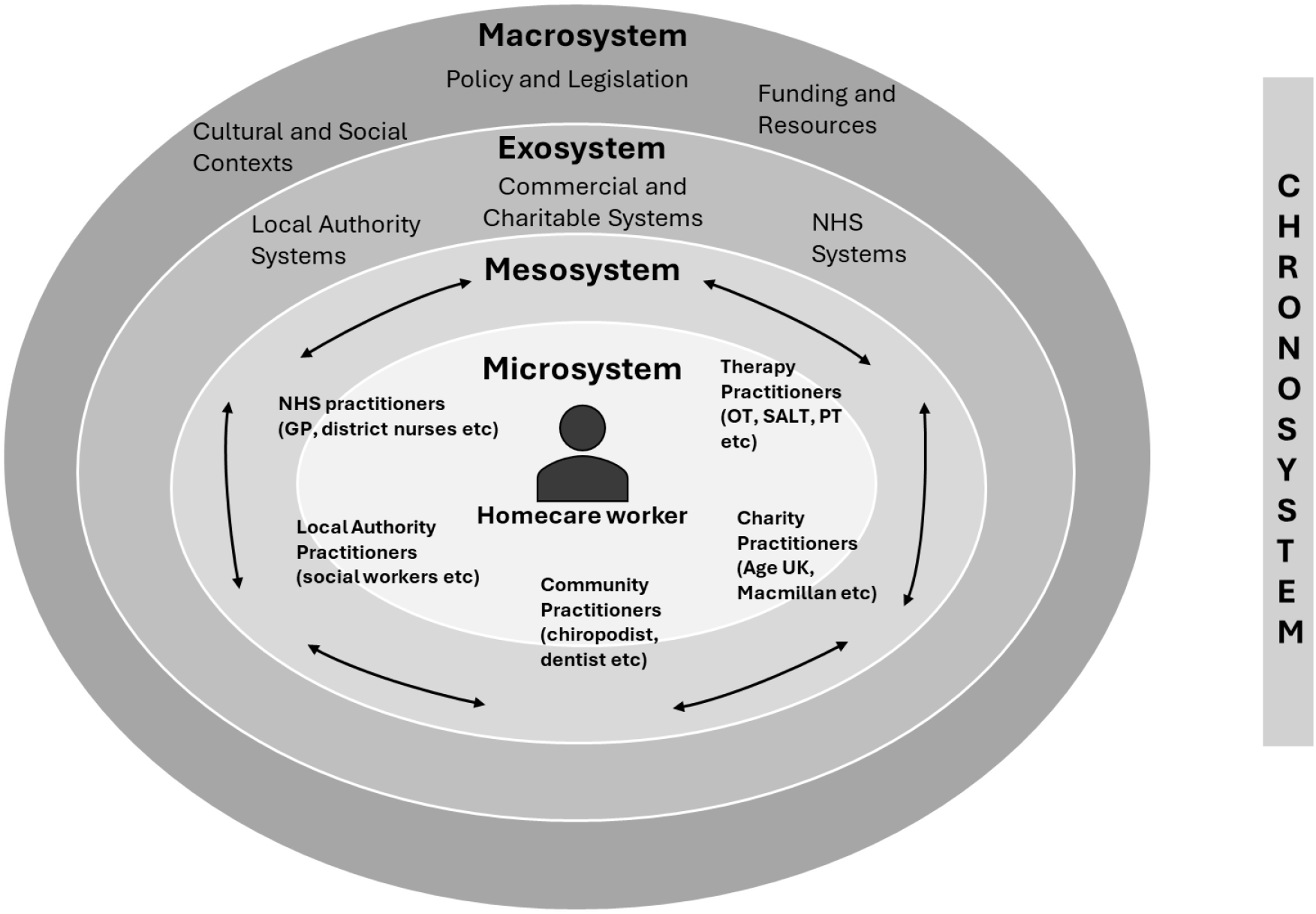
Ecosystem of Palliative Care Homecare Workers

In this paper we identify two key sub-themes of interprofessional collaboration which reflect the interaction between homecare workers and other practitioners: i) knowledge of the role of the homecare worker, and ii) valuing the homecare worker. Although there were good examples of collaborative working, most of our data identified barriers to effective collaboration (see Figure 2), which were seen in all areas of the ecosystem.

**Figure 2:**
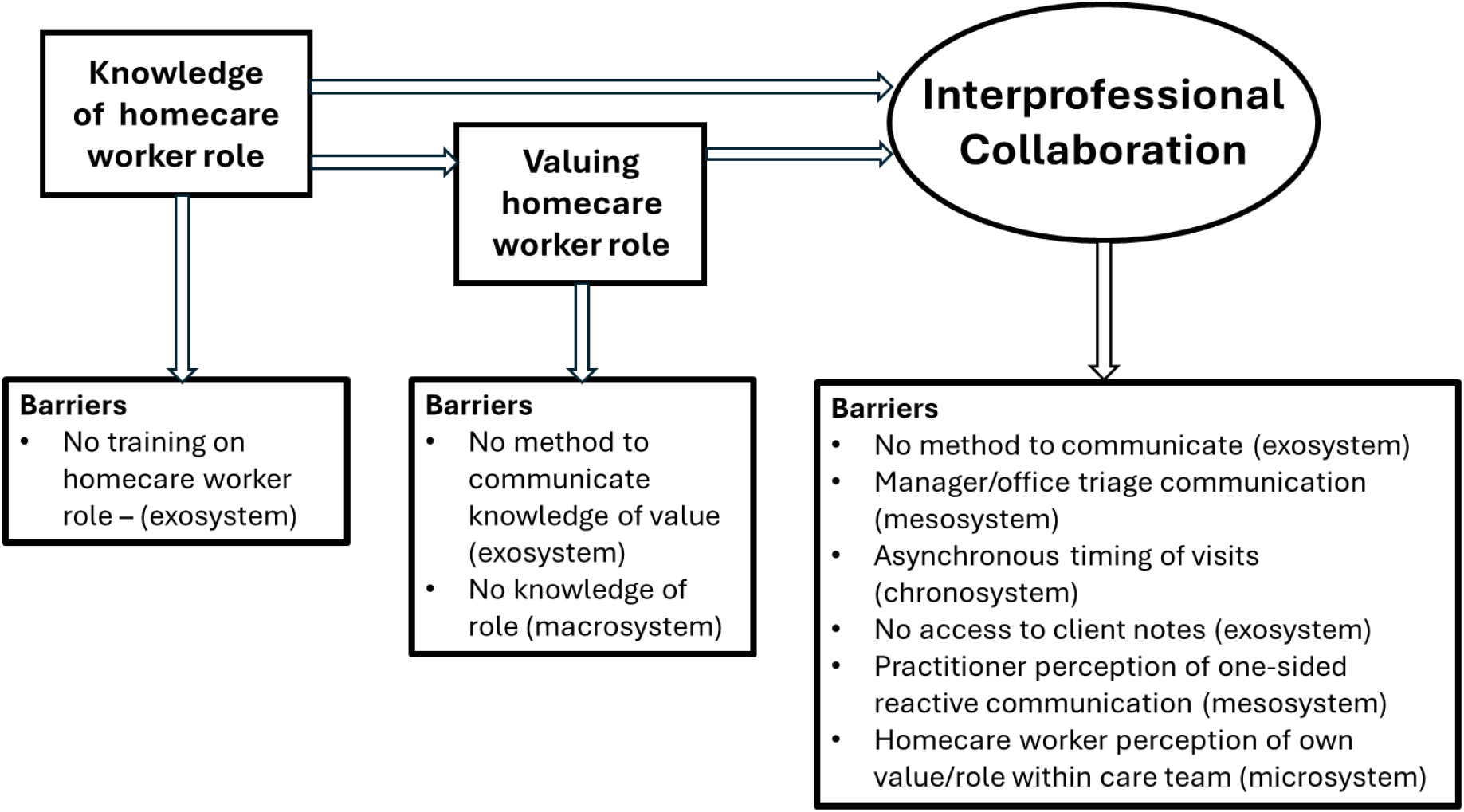
Enablers and Barriers to Interprofessional Collaboration and Connection to Relevant Ecosystem

### Knowledge of homecare worker role

Practitioner knowledge and understanding of the role of homecare workers varied. Some practitioners did not fully understand what homecare workers could and could not do in their role, causing difficulties with interprofessional working. There was no understanding of how to develop this knowledge, or how it could improve care.

Homecare workers and managers perceived practitioners as showing little recognition of the need to include homecare workers in interprofessional interactions, highlighting a lack of knowledge of the benefits of their role to people receiving care and the wider palliative care community.

> *“If you need a plaster and stuff you’d have to ring (laughs) a district nurse. So they kinda like get annoyed that they have to come out for such little things, but I don’t think they understand the rules of the homecare, that we aren’t allowed.” 3-HOMECARE WORKER -17*
>
> *“We are the ones that have the most contact, we are the ones that have that information, they don’t read our notes, they, they’re not interested in what we do, they’re not interested, yet if they were they would know and understand that person that’s lying in that bed much better than what they do.” 1-MANAGER-2*

### (De) Valuing the homecare worker role

Similarly, there was an underlying need for respect and trust, and valuing of the homecare worker role. Valuing their role could empower homecare workers to interact with practitioners on an equal level, and for practitioners to adopt a supportive and collaborative ethos. Although there was evidence that homecare workers were appreciated by some practitioners, there was uncertainty around how, or if, to share that view with homecare workers. Our study also evidenced some expressions of devaluing and lack of respect for the role of homecare worker by other practitioners.

> *“I would love to tell each individual care provider how valuable I find them, but I also am not sure that it’s just my responsibility to do that, or healthcare professionals’ responsibility to do that.” 1-PRACTITIONER-4*
>
> *“It feels as though caring is a job that people go to when they can’t do any other job.” 2-PRACTITIONER-3*
>
> *“They* [other practitioners] *don’t listen, they put their nose up a lot* [to show rejection of something that’s unworthy] *of the time to the carers, these nurses and that” 2-HOMECARE WORKER -3*

This was perceived to impact on practice, with practitioners and homecare workers working separately and without mutual support. The lack of understanding and appreciation of roles created and exacerbated interprofessional divides and poor communication.

> *“We have had district nurse teams where the carer has been there and incontinence has happened but the district nurse will refuse to assist the carer to help the person because that’s not their job to do personal care, they’ll arrive and they change a dressing, but they’ve been incontinent they won’t change the dressing until we send another member of staff to assist with personal care. So, it creates a real divide.” 3-MANAGER-2*

Achieving effective collaboration therefore between practitioners and homecare workers requires knowledge and valuing of the role, as evidenced in the sub-themes. However, other barriers adversely affecting collaborative practice were seen, beyond those associated directly with our sub-themes.

### Barriers to interprofessional collaboration

Direct communication with homecare workers was noted as a key challenge. Many agencies managed all communication centrally through office staff or managers, rather than homecare workers talking directly to practitioners. This was often justified as ‘protecting’ the homecare workers so they can continue working, and a practical solution due to limited time available for phone calls.

> *“We’ve always said if they’ve got any concerns… or they think anything could be changed, then to speak to us and we’ll do the phone calls for them.” 1-HOMECARE WORKER -9*

This, and the lack of synchronous working can be frustrating for all care providers as it impacts on direct communication, and opportunities for supporting homecare workers by providing valuable *ad hoc* training. This situation also limited the valuable contribution homecare workers could make to clinical care and planning, given their daily contact.

> *“I think if we could look at a way to try and (sighs) have more joint visits with carers there, and if we could maybe give carers some confidence in end of life care and to be able to speak to us and raise a few more things than they do; I think probably [we] maybe need something where we approach them as well, I think.” 3-PRACTITIONER-3*

Trying to arrange joint visits proved problematic due to different approaches to scheduling. Homecare workers often have no set visit times. Similarly, other healthcare practitioners such as district nurses and General Practitioners (family doctors) would visit as part of a scheduled round of calls, making it impractical to coordinate simultaneous visits.

> *“They don’t have time to sit for the community nurse to get there… sometimes their care calls are only half an hour/twenty minutes.” 3-PRACTITIONER-8*

One homecare agency illustrated highly effective interprofessional working relationship with practitioners, with regular communication, meetings, sharing expertise, resources, and support.

> *“The system that we use is used by all the nurses, all the admins, all the care, you know, our staff, we all access the same details, and you’ve also got physios that have been and documented notes and then you’ve got information about equipment or profiling beds that have been put in, cos we’re provided with them.” 3-HOMECARE WORKER -5*

This was enabled by the agency being sited within a statutory healthcare provider, which was an exception within our sample. Most homecare workers were employed outside the health authority and within private organisations, often subcontracted by local authorities. Access to, and communicating through, patient records was thus not possible. Both homecare workers and practitioners noted how difficult it was to access and share patient information:

> *“We’ve* [healthcare practitioners] *got a system where we can look behind the scenes and kind of see if a speech and language therapist has been in or an OT* [occupational therapist]; *we can have a little look behind the scenes to see their assessment, which is really helpful*… *For carers going in I suppose it’s a little bit more feeling in the dark because they’re not seeing that assessment, they’re just hearing it from the family, and sometimes families distort things to how they want it to be, or they forget things, or they can’t feed it back. So, the carers don’t quite get relayed the information that we can.” 2-PRACTITIONER-1*

When homecare workers were able to interact with other practitioners, there were significant benefits for those involved. Homecare workers commented on receiving support and training to help provide aspects of care, for example, how to move people safely, use equipment, or manage specific needs such as catheters:

> *“We would discuss with carers if they were there at the visit anything that maybe they weren’t doing or they needed extra support with, we would support the carers with that as well, like teaching and things like that, like how to use the slide sheet correctly, catheter care, anything else that basically they were unsure about.” 3-PRACTITIONER-8*

Our data indicated that practitioners often felt their relationships with homecare workers were reactive. They perceived it was the homecare workers’ responsibility to initiate contact for specific needs or when requiring assistance from practitioners, indicating a perception they were peripheral to the palliative care team.

> *“We’ve already got lots of people to speak to on a daily basis and teams and services, so, you know unless there’s a need for it, we probably wouldn’t be going to reach out to the care teams, but we’re always there if ever they wanted to speak to us as well, you know, if they needed to.”1-PRACTITIONER-3*
>
> *“If I wanted to give them a heads up about something I could just phone their care manager who would then pass the message onto the carers” 2-PRACTITIONER-9*

A two-way flow of communication was perceived by practitioners to be needed for effective collaboration. Both homecare workers and practitioners experienced frustration at not being able to talk directly, and therefore potentially not receiving enough information.

> *“When you ring the care company it’s not actually the carer you’re speaking to, you’re speaking to a manager… so it’s difficult to even speak to the actual person that raised the concerns, for example, and sometimes they might handover to somebody else, especially if they’re going off shift, sometimes they don’t, so even that person might be wondering why was this call made and I have no clue.” 1-PRACTITIONER-5*

Evidence of the challenges in relation to interprofessional collaboration was also found in the Pictor diagrams (see Figure 3 for an illustrative example). While many different practitioners were recalled as being a part of the microsystem around the person receiving care, homecare worker interactions focused on the client, their family, and the care agency. Even qualified social work professionals did not appear to have direct contact with homecare workers, highlighting their isolation and a lack of support from other practitioners.

**Figure 3:**
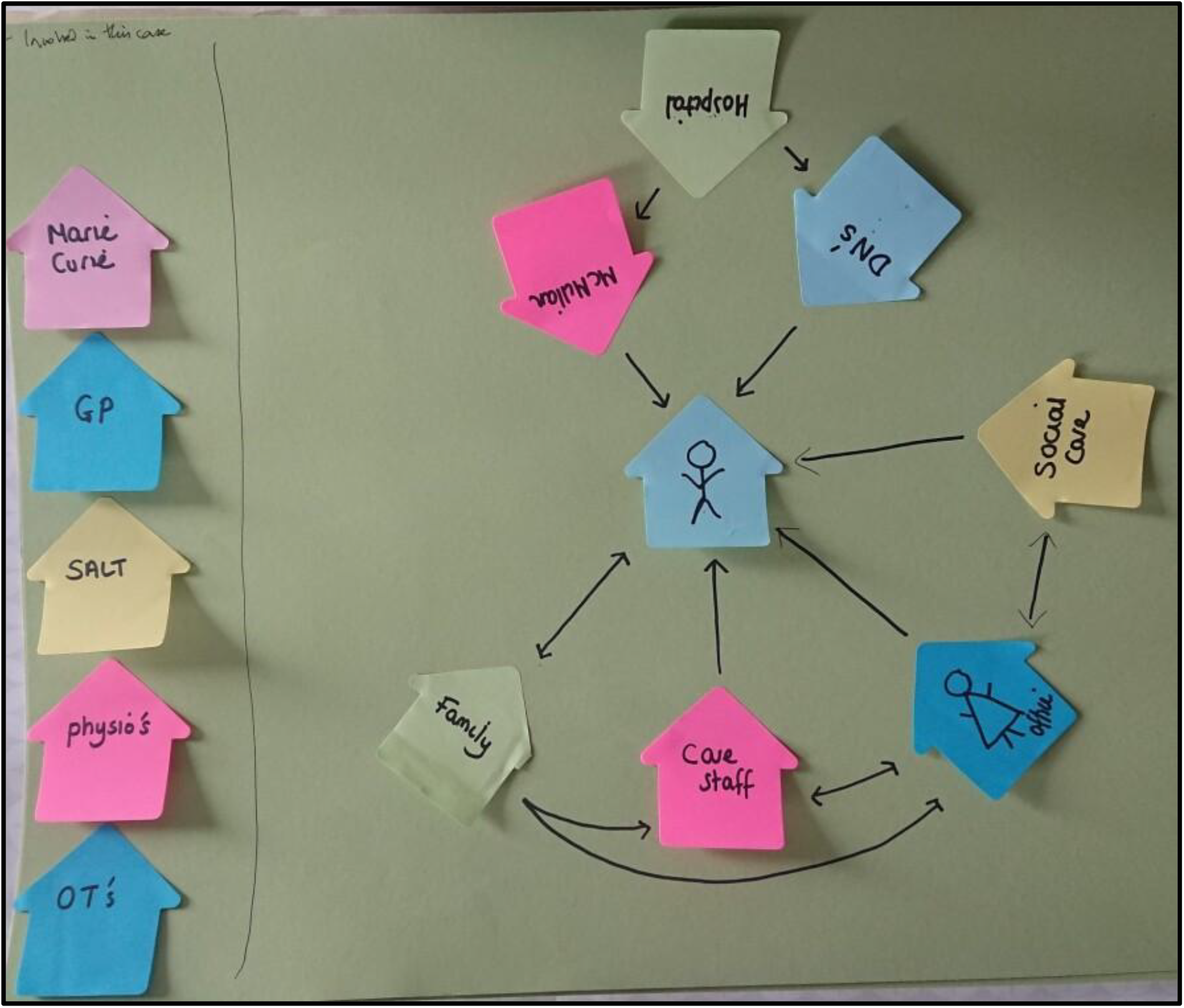
PICTOR 3-MAN-6. The ‘stick person’ at the centre, represents the person receiving care. SALT – speech and language therapist; GP – General Practitioner; DN – district nurse; MacMillan – palliative care nurse specialist; Marie Curie-hospice charity; OT’s – occupational therapist

## Discussion

### Main findings

We found that effective interprofessional working could be challenging, impacting all environmental levels. Despite good practice examples, the homecare worker role within end-of-life care appears to be poorly understood and under-valued by other practitioners, and the sometimes the homecare workers themselves. Inadequate collaboration caused feelings of isolation and eroded mutual trust. Barriers to effective interprofessional working included lack of training on the homecare role and its value for practitioners, lack of direct communication systems, gatekeeping of communication by managers, asynchronous working practices, misperception of reactive one-sided communication, and restricted access to respective documentation. One of the strongest facilitators was when the homecare service ‘sat’ within the statutory service provider enabling direct communication and access to joint documentation; however, this was unusual. Interaction between homecare workers and other practitioners enhanced respect, mutual learning, and enabled homecare workers to contribute their knowledge of the person receiving care to other practitioners.

### What this study adds

Our findings are consistent with other literature showing homecare worker isolation and eroded mutual trust caused by ineffective collaboration^20,21^. This exacerbated homecare workers’ own difficulties in recognising their value within end-of-life care^22^. Worldwide, evidence shows that collaborative practice improves quality assessment, compliance and patient satisfaction^23^. It is seen as integral to palliative care^1^. Conversely, poor practice reduces quality of care, and care recipient experience^24^.

We build on this literature by demonstrating that despite pockets of good practice, the dominant narrative is one of homecare worker isolation, with scarce direct contact with other practitioners. We describe a lack of knowledge of the homecare worker role leading to undervaluing (by practitioners *and* homecare workers), exacerbated by poor understanding of the contribution that homecare workers bring. Healthcare practitioners need to be knowledgeable about the roles of *all* those working with people approaching end-of-life, and *vice versa*^25-27^ for effective interprofessional working. In our data, even when some practitioners stated they valued and respected homecare workers, they felt unable to express this because there was no process to do so, often due to workplace systems and organisational cultures^24^.

We demonstrated little interaction between practitioners and homecare workers, due to the triaging of communication by homecare agency managers or office staff and a lack of supportive structures and systems. Reliance on one form of communication, e.g., telephone, negatively affects working relationships^21^, and poor communication has often been noted within palliative care^28^. We evidenced that other forms of communication (e.g., electronic clinical records) can be a barrier^29^; a particular challenge for homecare workers who sit outside health and social care structures, often with separate systems. One connected communication system, as an exosystem change, could enable greater flow of information and communication between the extended multi-disciplinary team^30^. The use of everyone’s expertise and experience is a key component to providing holistic palliative care^22^. However, we showed a reliance on reactive communication by practitioners. Interprofessional communication and collaboration fosters a sense of security for care recipients and family carers^31^. A communication block from homecare worker to healthcare practitioners leaves a clinically important gap.

There were barriers to interprofessional working within the chronosystem due to the asynchronous timing of practitioner visits. Homecare workers were not aware of, or able to be often present when other practitioners provided care and *vice versa*. Where this did occur, it enabled reciprocal communication and collaborative working, including training opportunities. The lack of integration of resources such as time has been noted within palliative care^24^ and can be an issue at institutional and structural levels within the exosystem due to working practices, impacting on mesosystem interactions between practitioners.

The World Health Organisation identified education as a crucial component of palliative care provision^32^. We argue this should be a two-way process between health and social care, including homecare workers^33^. The need for education impacts on the meso-, macro- and exo-systems in relation to accessing appropriate training within the workforce. Within society there is a need for wider recognition of the importance and value of the role of the homecare worker, and their inclusion within key documentation and policy where they have been overlooked in the context of end-of-life care.

### Strengths and limitations

We had good representation across all participant categories, within three diverse regions in England, with rich data providing sufficient information power to address our research questions. Our analysis included using our public contributors to sense check the findings, and the interpretation of themes. Although only a third of participants created a Pictor diagram; this nevertheless provided a helpful complement and sense-check of interview findings.

We acknowledge the structure of community-based end-of-life care will be different within England to other countries, limiting the relevance of our findings to other areas that have more integration between health and social care workforces. However, we propose that our findings are applicable not only to other UK regions but may also highlight potential interprofessional working barriers in social and health care models internationally, based on the existing evidence. We offer robust insight into homecare provision, an under-researched and timely topic given the current crisis in homecare provision in the UK^35^.

### Recommendations for practice and policy

We raise key challenges for those commissioning and providing end-of-life care services. There is little research around the crucial role of homecare workers supporting people to live at home when approaching end-of-life^36^. We highlight that role in the context of collaboration and interprofessional working; there is a need for practitioner education around the homecare worker role and their value in community-based care. We also suggest consideration of wider institutional and organisational changes including timing of visits, sharing of records, and direct two-way communication streams. This would require transformation across all systems and organisations providing care^24,30^ to empower homecare workers as an integral part of end-of-life care provision^32^. This will include a cultural change to enable collaboration and interprofessional working^22^. This change needs to be across the whole ecosystem, incorporating different systems and levels of care within the health sector, social care, community support and wider societal networks^29^.

Our findings could be applicable to all home-based care, as many of the barriers identified are not unique to end-of-life, or to England. Issues around interprofessional collaboration may have broader implications for community-based care within people’s homes.

Finally, a wider societal acknowledgement, including policy, of the value of homecare workers is long overdue^20^. This poorly understood and under-researched group enable people to be supported in their preferred place of care, and place of death; the significance of their contribution should not be underestimated.

## Conclusion

The homecare worker role is often poorly understood, undervalued and inadequately communicated. Effective collaborative social and healthcare practice is necessary for high quality care but was often lacking in our findings which should challenge community end-of-life care providers. Mutual education of roles, and facilitation of direct contact between homecare workers and other practitioners helps address shortfalls. However, definitive improvement will need far-reaching organisational, political and societal changes to overcome serious barriers resulting from different working structures. Further research should explore strategies to address the barriers identified.

## Data Availability

All interview data in the present study are available upon request to the authors.
https://hull-repository.worktribe.com/output/5179695.

## Author contributions

ZB drafted article. All authors read, commented on, and approved the final manuscript. Conceptual design of study MJJ, LW, AB, PT, MP, JW and CW. Data collection and analysis ZB, CF, JK, HE-B, AB, CW, LW, MJJ.

## Availability of data and materials

The data is not available publicly due to the sensitive nature of content.

## Acknowledgements

The authors would like to thank the participants of the study who generously shared their time and expertise with the research team. We would also like to thank the wider research team including Kathryn Harvey of Hull York Medical School, and Joan Bothma of Cera Care, and the people with lived experience who contributed to the research.

