## Supplemental Table A for "Addressing Barriers to Interprofessional Working with Homecare Workers in Community Palliative Care: Insights from a Multi-site Qualitative Case Study in England"

UNIVERSITY
OF HULL

**Rapport-building questions:**

Can you tell me how long you've been working in social care or homecare?

How long have you been working for [name of agency]?

**Questions about the care they have provided to a client**

The Pictor technique will be introduced to help the interviewee think about someone (a client) they currently or recently have supported in the last months, weeks or days of life. The technique focuses on roles and relationships – when interviewees are asked to 'tell the story' of the Pictor chart they have produced, they are likely to spontaneously cover the following issues, with minimal probing questions from the researcher, using the chart as a prompt:

- What kinds of care did you provide as part of the support plan?
- Was there an unpaid carer (such as a family member or friend)? Did you provide any support to them? Did they live with the person or somewhere else?
- Who else was involved in the care and treatment of the client?
- How did you get on working with those other people? How did those other people work with you/treat you? Did they involve you in any discussions or decisions about the person's care?
- What support did you get from your colleagues/managers? Was there any support you needed but didn't receive?
- Did you experience any difficulties or dilemmas when supporting the person? Did you experience any difficulties or dilemmas when working with carers? Did you experience any difficulties or dilemmas when working with other professionals? How did you manage/resolve these things?

If the participant does not want to make a Pictor chart, they will be asked to think about a client they have supported recently/currently, and will be asked the same set of questions.

**Questions about providing care to someone in the last months, weeks or days of life:**

What do you think are the main differences in providing homecare at this time of life, compared to providing care to clients who are not at the last stage of their illness?

What are the main challenges in providing support to people when they are dying at home (when they are in the last months, weeks or days of life? Are there any 'positives' in this? What are the main things which help you when providing care in this situation?

What are the main challenges in providing support to carers (families and friends) when someone is dying at home? What are the main things which help you?

How are you affected emotionally when someone is dying? Who or what helps you at this time?

How are you affected emotionally when someone dies? Who or what helps you at this time?

Do you ever have contact with the person's family/friends after they have died?

**Questions about training**

Have you received any training in caring for people at the end of life? Who provided this training?

Has your agency done anything to help prepare you for working with people at the end of life?

Is there any training you would like to help you in any aspect of caring for people at the end of life?

Do you have any friends or colleagues working as homecare workers who might be interested in taking part in this research? Would you be willing to pass on some information about the research to them?
