## Supplemental Table B for "Addressing Barriers to Interprofessional Working with Homecare Workers in Community Palliative Care: Insights from a Multi-site Qualitative Case Study in England"

### SUPPORTED FINAL THEMES

Themes used within this paper are highlighted:

| | |
|---|---|
| **Theme 1 - End-of-life care presents unique privileges and challenges** | |
| ***A*** | *Homecare at EoL requires flexibility to respond to changing needs and circumstances* |
| | Client-HCW communication changes<br>HCWs are required to work flexibly<br>Managing deterioration and change<br>Timing and flexibility of support when calls run over |
| ***B*** | *EoLC is experienced as a privilege but makes unique demands and impacts on HCWs* |
| | EoLC is experienced as rewarding<br>HCWs are committed to providing good EoLC<br>Care may be set up as EoLC or long-term care which becomes EoLC<br>Emotional impact of anticipating the loss - being present at death<br>EoLC is distinct from other homecare - and may be more challenging<br>EoLC present specific uncertainties<br>Funerals and staying in touch with families after a death<br>HCW confidence in speaking about death - dying<br>HCWs experience bereavement and loss<br>Practical tasks when someone dies<br>Specific tasks are required<br>Wear many hats |
| ***D*** | *HCWs require specific support when providing EoLC* |
| | HCW managing own emotions incl resilience<br>Recognition that this is not for everybody<br>Support from managers<br>Who supports the HCW |
| **Theme 2 - EoLC is provided in a relational and personalised context** | |
| ***A*** | *EOLC is delivered in response to individuals' personal and holistic needs* |
| | Continuity and consistency of care<br>Issues relating to personal and intimate care<br>Personalities and prejudices inform care experience<br>Support for gender, culture, ethnicity<br>Support for social needs<br>Support to live while dying<br>Trust, dignity and respect |
| ***B*** | *HCW role at EoL is carried out in a relational and family context* |
| | Caring together - HCW and family provide care<br>Contact with non-resident families<br>HCWs advocate for client<br>Importance of relationship with client at EoL<br>Professional spends most time with client<br>Supporting family not on care plan<br>Supporting the whole family |
| ***C*** | *HCWs need to negotiate relationships and boundaries* |
| | Being an intermediary or stand-in for family |

| | |
|---|---|
| | Boundaries in relationships<br>Expected to care but not too much<br>Family understanding and expectation of HCW role<br>Managing family dynamics and conflicts<br>Rapport with clients and families<br>Whose wishes and priorities inform care |
| ***D*** | *HCWs support and navigate other people's emotions and 'read the room'.* |
| | Emotional support is not always commissioned but important<br>Responding to other people's emotions<br>Using intuition and reading the room |
| **Theme 3 - Homecare at EoL takes place in context of multi-agency teams and hierarchies** | |
| ***A*** | *Importance of Interprofessional Collaboration and effective working relationship* |
| | Communication between HCW and practitioners<br>Communication between HCWs, colleagues and managers<br>Importance of effective relationships<br>Tasks and boundaries - what belongs to HCW – HCP<br>Access to practitioners<br>Managers as gatekeepers<br>Documentation quality |
| *B* | *Interprofessional working can marginalise and isolate HCWs* |
| | HCW could do more<br>HCW isolated and separate<br>Perceived value and trust of HCW by others<br>Perception of simplicity of tasks<br>Status of HCWs - just a HCW<br>Health records and systemts including access<br>Paper-app notes |
| **Theme 4 - HCWs have specific knowledge and training needs at EoL** | |
| ***A*** | *Preparedness* |
| | Do they know they will be providing EoLC at outset<br>Knowledge on what to do at time of death |
| ***B*** | *Access and availability of training* |
| | Access to EoLC training<br>Time for training |
| ***C*** | *Identified training needs and gaps* |
| | Basic care (not EoL) |
| | Accessing Support |
| | Symptom Management |
| | Palliative v EoL v 'normal' care – what to expect/trajectories/shifts in priorities |
| | Specific tasks more common at EoL |
| | Emotional and psychological aspects |
| | Religion, culture, and death |
| | Communication |
| ***D*** | *Sources of knowledge* |
| | HCW bring experience of EoLC from other settings<br>HCWs can be reluctant to say they don't know<br>Learning is through experience and 'on-the-job' in the workplace<br>Reliance on HCW as 'natural' carer rather than formal training |
| **Theme 5 - External factors** | |

| | |
|---|---|
| ***A*** | *Operational Factors* |
| | Expectation HCW to go beyond hours<br>Recruitment and retention<br>Where HC services are situated |
| ***B*** | *Variations in homecare agency working and quality* |
| | Different approaches to EoL care and tasks in different agencies<br>How different services are run<br>Lone working<br>Variation in quality of homecare (between agencies - HCWs) |
| ***C*** | *Regional differences in support* |
| | local - regional differences in external support<br>OOH delivery - support for OOH HCWs |
| ***D*** | *Challenges re funding and information* |
| | challenges re funding and information<br>Perception that funding for CoLC is insufficient |
